# Epidemiology of Venezuelan haemorrhagic fever in Barinas state, Venezuela

**DOI:** 10.64898/2026.04.04.26348925

**Authors:** María-Mercedes García, Xacdiel Rodríguez, Santiago José López, Jose Jose Reyes Dorante, Emilsa Jaqueline Aldana, Niyuris Elena Orduño, Aurora Lugo, Daniel Salazar, Nailyn Carvallo, Yismar Rivas, Cassia F. Estofolete, Mauricio Lacerda Nogueira, Carlos Lezcano-Coba, Josefrancisco Galué, Yelissa Juárez, Christl A. Donnelly, José Narciso Franco, Jean-Paul Carrera

**Affiliations:** Ministerio del Poder Popular para la Salud, Barinas, Venezuela; Carson Centre for Health and Ecosystem Research, Darien, Panama; Facultad de Salud Pública y administración, Universidad Peruana Cayetano Heredia, Lima, Perú; Faculdade de Medicina de São José do Rio Preto, São Paulo, Brazil; Departament of Pathology, University of Texas Medical Branch, Galveston, TX, USA; Department of Statistics and Pandemic Sciences Institute, University of Oxford, Oxford, United Kingdom

**Keywords:** Guanarito virus, New World Arenaviruses, American haemorrhagic fever, Zoonoses, Infectious diseases

## Abstract

**Background:** Venezuelan haemorrhagic fever (VHF), caused by Guanarito virus (GTOV), is a zoonotic disease endemic to the western plains of Venezuela. Despite decades of recognition, its epidemiology and clinical profile remain poorly characterised.

**Methodology:** We analysed individual-level data from standardised case report forms submitted to the Venezuelan National Epidemiological Surveillance System between 2017 and 2024 for suspected VHF cases in Barinas, Apure, and Portuguesa. Demographic, clinical, and laboratory variables were examined to characterise temporal and geographical patterns and to define the clinical profile of VHF compared with endemic arboviral infections.

**Principal Findings:** Among 480 suspected cases, 72 (15.0%) were laboratory-confirmed GTOV infections. Confirmed cases occurred predominantly in men engaged in agricultural or service-related occupations, with the highest prevalence among individuals aged 46–90 years. A marked seasonal pattern was observed, with most cases occurring between September and January. The most frequently reported symptoms included headache, haemorrhage, sore throat, and diarrhoea. Compared with other endemic arboviral infections, GTOV was more strongly associated with headache, myalgia, sore throat, haemorrhage, and abdominal pain, delineating a distinct clinical phenotype relative to diseases caused by encephalitic alphaviruses, chikungunya virus, dengue virus, and Zika virus. The case fatality ratio among laboratory-confirmed cases was 36.1% (95% CI: 25.1–48.3). GTOV infection was independently associated with mortality (adjusted relative risk [aRR] 3.66; 95% CI 2.28–5.87; *p* < 0.001), underscoring its substantial clinical severity.

**Conclusion:** GTOV remains endemically transmitted in western Venezuela, disproportionately affecting older men engaged in agricultural and service-related occupations. Its seasonality and clinical phenotype, characterised by haemorrhage, sore throat, and gastrointestinal symptoms, highlight the need for clinical awareness and improved differential diagnosis, particularly in remote endemic settings with limited access to laboratory testing.

**Author Summary:** Venezuelan haemorrhagic fever is a serious disease with poorly understood epidemiology and clinical presentation and affects rural communities in western Venezuela. Although it has been recognised for decades, important gaps remain in understanding who is most at risk, how the disease presents, and why it leads to high mortality. In this study, we analysed national surveillance data collected between 2017 and 2024 to better describe its epidemiology and clinical features. We found that the disease mainly affects older men involved in agricultural and service-related work, with cases occurring more frequently in September and January. Patients commonly presented with headache, haemorrhage, sore throat, and gastrointestinal symptoms, forming a consistent set of clinical patterns. Infection was strongly associated with an increased risk of death. These findings highlight the need for earlier clinical recognition and improved diagnosis, particularly in resource-limited settings, and support the development of targeted public health actions to reduce preventable deaths in high-risk populations.

## Introduction

Venezuelan haemorrhagic fever (VHF) is a zoonotic viral disease caused by Guanarito virus (GTOV), an arenavirus related to Lassa virus, the causative agent of Lassa fever [1]. GTOV is a New World mammarenavirus and is more closely related to South American haemorrhagic arenaviruses, including Junín virus, Machupo virus, and Chapare virus [2,3]. GTOV is maintained in a sylvatic cycle primarily involving rodent reservoirs, including the short-tailed cane mouse (*Zygodontomys brevicauda*), with human infection occurring through contact with or inhalation of contaminated rodent excreta and bodily fluids [4,5]. Since its initial identification during a severe outbreak in Guanarito, Portuguesa State, in 1989, VHF has persisted as an endemic zoonosis in rural agricultural regions of western and central Venezuela, particularly in the states of Portuguesa and Barinas, with ongoing sporadic transmission documented through national surveillance [6].

Clinically, VHF typically presents as a non-specific febrile illness that may progress to severe haemorrhagic manifestations, complicating early differentiation from other endemic infections, including severe dengue and co-circulating arboviral diseases [7]. However, detailed comparative analyses of these co-endemic infections remain limited. This clinical overlap hinders differential diagnosis, particularly in resource-constrained settings where the burden of undifferentiated febrile illness is high.

Although cases have been reported since the late 1980s, much of the available literature has focused on the initial outbreaks, with limited contemporary data describing clinical characteristics, seasonal dynamics, case fatality ratios, and associated risk factors [6,7]. Ongoing socio-environmental changes, including agricultural expansion, climate change, and increased human mobility, are likely influencing the transmission dynamics and geographic distribution of GTOV [8]. To address these knowledge gaps, we conducted a retrospective analysis of hospital-based surveillance data collected between 2017 and 2024 from the National Epidemiological Surveillance System to characterise the clinical presentation, epidemiological patterns, case fatality ratio, and risk factors associated with VHF in western Venezuela.

## Methods

### Study design

We conducted a retrospective study using data collected between 2017 and from the Venezuelan National Epidemiological Surveillance System. The system, administered by the Venezuelan Ministry of Health, uses standardised case report forms to collect demographic, clinical, epidemiological, and laboratory information on suspected and laboratory-confirmed VHF cases. Most notifications originated from public hospitals in the western plains, particularly Barinas State, a recognised endemic area for GTOV circulation (Fig 1A).

**Fig 1.**
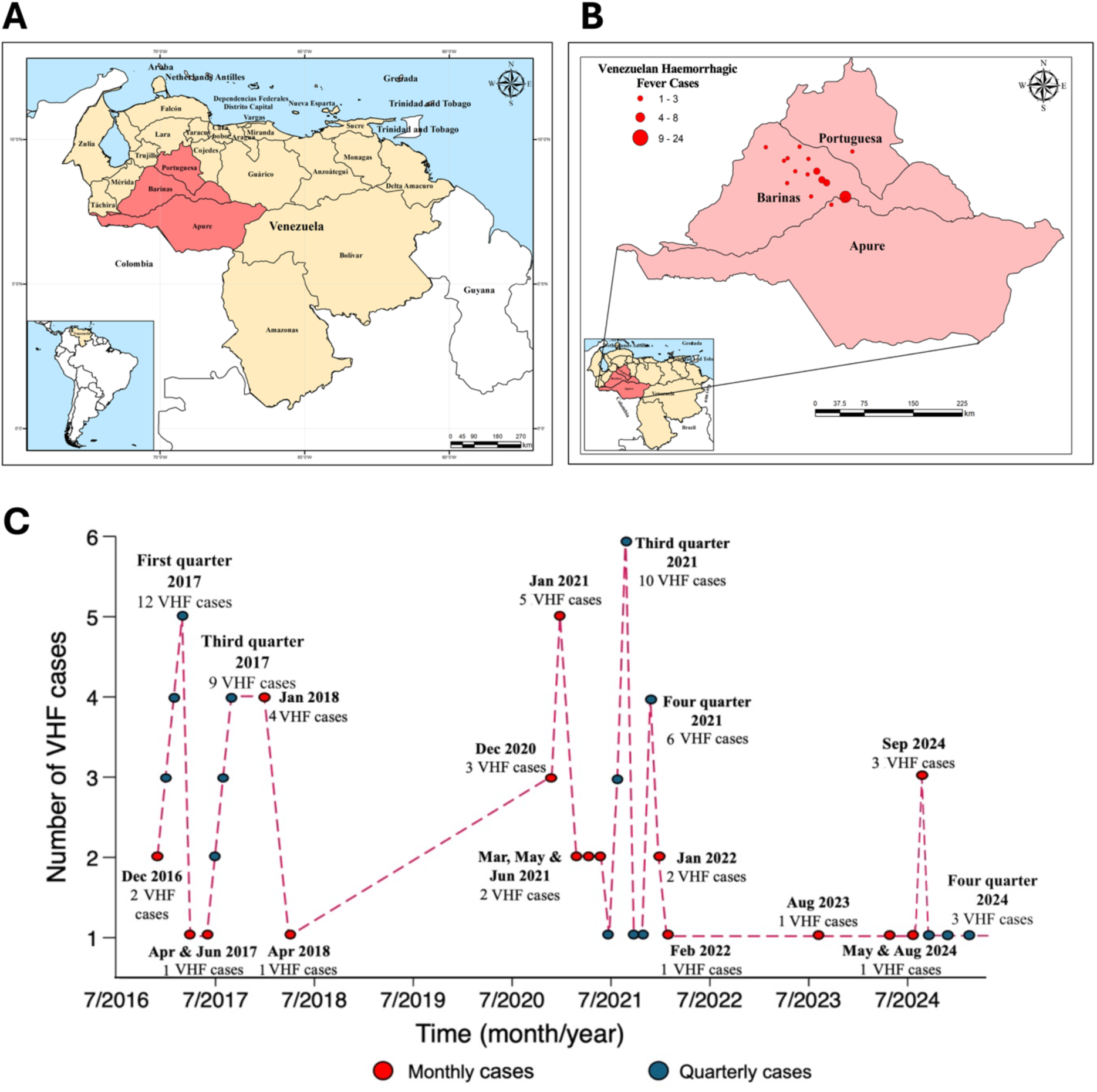
Study area and spatio-temporal patterns of confirmed Venezuelan haemorrhagic fever cases in Venezuela (2017–2024). (A) Map of Venezuela showing the study area, which includes three states located in the western plains region: Barinas, Apure, and Portuguesa. (B) Geographic distribution of confirmed VHF cases across municipalities within the study area, the red circle size on the map reflects the number of reported cases per locality. (C) Temporal distribution of confirmed VHF cases. In the time series, red dots represent the number of confirmed cases reported in each individual month. Blue dots correspond to the cumulative total for the respective calendar quarter. Although plotted at monthly intervals for visual alignment, the blue values reflect aggregated quarterly totals rather than individual monthly counts.

Eligible records corresponded to individuals meeting national surveillance definitions for suspected or probable VHF with available laboratory test results. Case classifications were based on established national surveillance criteria (S5 Table) [9,10]. To minimise outcome misclassification, records with laboratory-confirmed alternative diagnoses of viral or bacterial origin were excluded from the analysis.

### Laboratory diagnosis

Clinical specimens—including whole blood, serum, and tissue samples—were collected at healthcare facilities and transported under cold-chain conditions to the Rafael Rangel National Institute of Hygiene, the national reference laboratory for VHF diagnostics. Laboratory confirmation was performed using enzyme-linked immunosorbent assays (ELISA) to detect IgM and IgG antibodies against viral haemorrhagic fever pathogens, as well as polymerase chain reaction (PCR) assays for the detection of viral RNA [11,12]. All diagnostic procedures were conducted in accordance with standardised national laboratory protocols [13].

### Comparison of GTOV symptoms with endemic arboviruses

To contextualise the clinical profile of VHF, we compared laboratory-confirmed GTOV cases reported in Barinas state between 2017 and 2024 with cases reported to other endemic arboviruses, including encephalitic alphaviruses—specifically Madariaga virus (MADV) and Venezuelan equine encephalitis virus (VEEV)—as well as chikungunya virus (CHIKV), dengue virus (DENV), and Zika virus (ZIKV).

Data for encephalitic alphaviruses (MADV and VEEV) infections were derived from Panama’s national surveillance system (1961–2023) and have been described previously [14]. Data on CHIKV infection were obtained from surveillance studies conducted between 2015 and 2020 in Amazonas State and Recife, Pernambuco, Brazil [15]. DENV and ZIKV datasets, from 2009 and 2016 respectively, were obtained from the São José do Rio Preto Health Service in São Paulo State, Brazil, and have been previously published [16].

### Statistical analysis

Descriptive analyses were performed to characterise demographic and clinical variables. Categorical variables were summarised using contingency tables and reported as absolute frequencies and percentages, while continuous variables (age) were described using the mean and standard deviation. The primary outcome for risk factor and symptom analyses was laboratory-confirmed GTOV infection, operationalised as a dichotomous variable (0 = suspected/probable case with negative molecular and serological results; 1 = confirmed case with positive PCR and/or IgM or IgG serology). Associations between categorical variables were assessed using Pearson’s χ² test, and Fisher’s exact test was used when expected cell counts were <5. Differences in mean age were evaluated using Student’s t-test.

Covariates for risk factor analyses were prespecified according to an epidemiological framework informed by prior literature and biological plausibility. These included sex, age (categorised into quintiles), occupation, and selected environmental and behavioural exposures, such as footwear use and the presence of rodents or crops near the household. For symptom-based analyses, variables were selected using a manual forward selection procedure based on comparisons of nested models. The final model was adjusted for sex and age (modelled as a continuous variable).

Generalised linear models (GLMs) with a Poisson distribution, a log-link function, and robust variance estimation were used to estimate relative risks (RRs) and 95% confidence intervals. This modelling approach was selected to provide stable and conservative effect estimates while minimising potential overestimation of associations. Statistical significance was defined as *p* < 0.05, and 95% confidence intervals were reported.

We performed comparative analyses to distinguish the clinical phenotype of confirmed GTOV infections from other endemic arboviral diseases in Venezuela, including encephalitic alphaviruses, CHIKV, DENV and ZIKV. To this end, four separate binary outcome variables were constructed, coded as 1 for GTOV and 0 for each comparator infection. Symptom variables exhibiting structural zeros, defined as the complete absence of observations within one outcome category were excluded to avoid model separation and non-convergence (S6 Table).

Variable selection was conducted using a manual forward selection strategy based on nested model comparisons, with candidate variables sequentially introduced and retained according to improvements in log-likelihood and statistical significance (*p* < 0.05). GLMs with a Poisson distribution, log-link function, and robust variance estimation were used to estimate adjusted relative risks and 95% confidence intervals. Sex and age were included a priori in all models as potential confounders, with age entered as a continuous variable. All analyses were performed using Stata version 18 (StataCorp, College Station, TX, USA).

### Ethical considerations

This study was conducted using anonymised data from Venezuela’s National Epidemiological Surveillance System. In accordance with national regulatory requirements and institutional policies, the study protocol underwent ethical and technical review by the Ministry of People’s Power for Health and the Institute of Advanced Studies “Dr. Arnoldo Gabaldón,” and received formal approval on 17 October 2025 (G-200006221-5).

## Results

### Demographic and epidemiological characteristics

From January 2017 to December 2024, a total of 719 cases of undifferentiated febrile illness compatible with VHF were reported to the Venezuelan National Epidemiological Surveillance System. All suspected cases were considered for inclusion. Of these, 59 individuals (8.2%) were excluded due to laboratory confirmation of alternative aetiologies. Viral infections accounted for 7.5% of excluded cases, including DENV (n = 49; 6.8%), yellow fever virus (n = 2; 0.3%), CHIKV (n = 1; 0.1%), and hepatitis A virus (n = 2; 0.3%). Bacterial infections comprised 0.7% of exclusions, including leptospirosis (n = 4; 0.6%) and rickettsiosis (n = 1; 0.1%). An additional 180 cases (25.0%) were excluded due to the absence of laboratory test results. The final analytical sample consisted of 480 participants, representing 66.8% of the initial cohort (S1 Fig).

Of the 480 suspected VHF cases included in the analysis, most cases were reported from Barinas (n = 464), with only a small number from Apure (n = 8) and Portuguesa (n = 8). Overall, 67.6% of participants were male and 32.4% were female, with a median age of 26 years (IQR: 16–41). Regarding occupational activity, 32.5% were engaged in agricultural or livestock-related work, 30.4% were associated with the educational sector (students and teachers), and 21.9% were economically inactive (including homemakers and infants). After stratifying by sex and restricting the analysis to adults (≥18 years), men were predominantly engaged in agricultural and livestock activities (67.1%), whereas most women were not economically active (82.9%). Most participants reported regularly wearing shoes (91.6%) and indicated the presence of rodents (95.8%) and adjacent crops (96.6%) in their residential environment (S1 Table).

### Prevalence and distribution of confirmed VHF cases

A total of 72 laboratory-confirmed cases of VHF were identified, corresponding to an overall prevalence of 15.0% (95% CI: 11.9–18.5). Infection was more frequent among males, with 16.7% (95% CI: 12.8–21.2) testing positive, compared with 11.6% (95% CI: 7.0–17.7) among females (Fig 2A). Age-stratified analyses revealed the highest proportion of confirmed cases in the oldest age quintile (46–90 years), with 20.0% testing positive, followed by the second quintile (14–22 years), with a prevalence of 15.5%. Prevalence varied by occupation, with the highest proportion observed among individuals working in the commercial and service sectors (21.5%; 14 cases), followed by those engaged in agricultural and livestock production (18.0%; 28 cases) (S2 Table).

**Fig 2.**
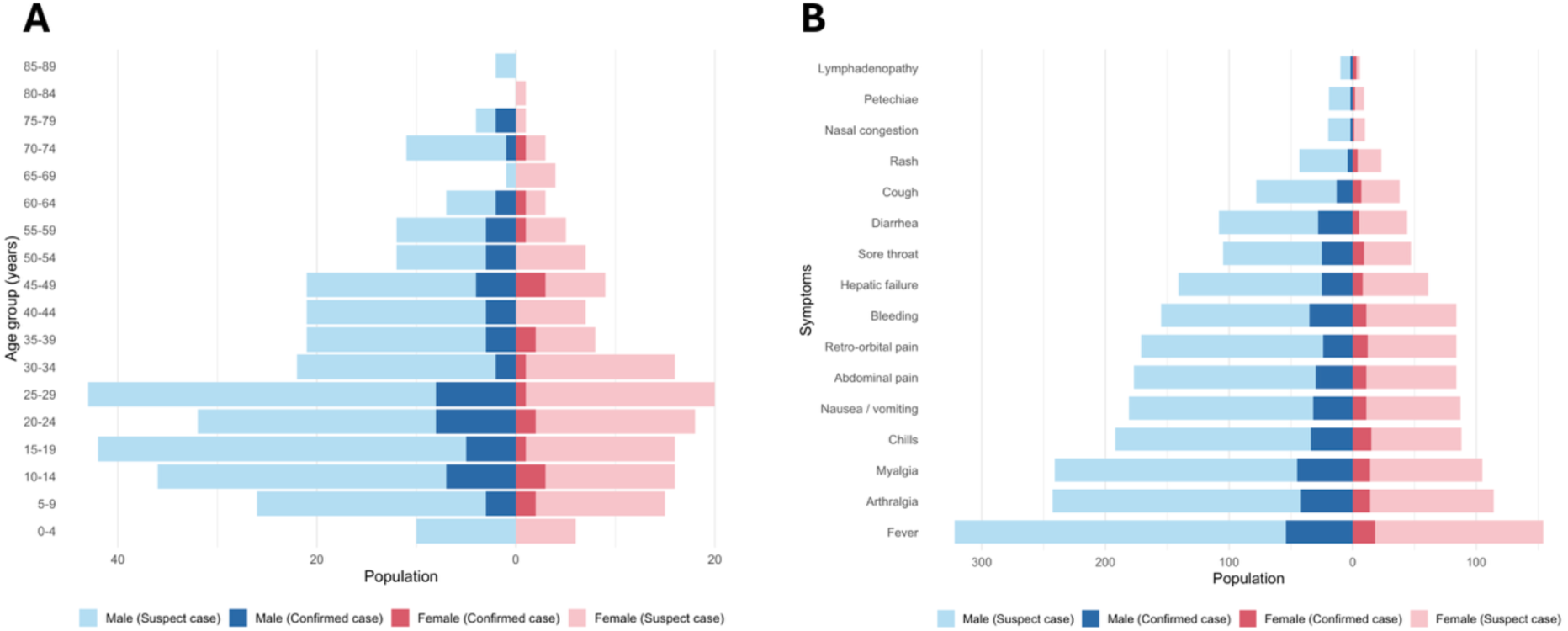
Nested population pyramids by sex, age group, and symptom profile. (A) Distribution of suspected and confirmed VHF cases by 5-year age groups and sex. (B) Distribution of suspected and confirmed VHF cases by symptom categories and sex. Male participants appear on the left and female participants on the right. Suspected cases are displayed in pastel tones, whereas confirmed VHF cases are shown in darker shades of the corresponding colours.

### Geographic distribution of VHF cases

Confirmed cases were identified across three states in the western plains of Venezuela, with most cases reported in Barinas State. In Barinas, 68 laboratory-confirmed cases were reported, corresponding to a prevalence of 14.7% (95% CI: 11.6–18.2). In Apure, three confirmed cases corresponded to a prevalence of 37.5% (95% CI: 8.5–75.5), and a single confirmed case in Portuguesa corresponded to a prevalence of 12.5% (95% CI: 0.3–52.7) (Fig 1B).

Within Barinas, cases were distributed across six municipalities: Sosa reported 25 cases (37.9%; 95% CI: 26.2–50.7), Rojas 23 cases (34.8%; 95% CI: 23.5–47.6), Obispos 9 cases (13.6%; 95% CI: 6.4–24.3), Barinas Municipality 7 cases (10.6%; 95% CI: 4.4–20.6), and Bolívar and Álvaro Arvelo Torrealba each reported one case (1.5%; 95% CI: 0.0–8.2). In Apure, all three cases occurred in Muñoz Municipality (2.9%; 95% CI: 0.4–10.1), and the single case in Portuguesa was reported in Portuguesa Municipality (1.4%; 95% CI: 0.0–7.8) (Fig 1B).

### Temporal patterns of infection

Regarding temporal distribution between 2017 and 2024, the highest number of confirmed cases occurred during the first (January-March: n = 26; 37.7%; 95% CI: 26.1–50.2) and third (July-September: n = 24; 34.8%; 95% CI: 23.7–47.2) quarter of the year. More than half of all confirmed cases (n = 38; 52.8%) were concentrated in three months—January, September, and December. January and September each accounted for 14 cases (19.4%; 95% CI: 11.1–30.5), followed by December with 10 cases (13.9%; 95% CI: 6.9–24.1) (Fig 1C).

### Clinical presentation of confirmed cases

The most frequently reported symptoms among laboratory-confirmed VHF cases were fever (100.0%), headache (95.8%), myalgia (82.0%), arthralgia (77.8%), chills (68.1%), haemorrhage (63.9%), nausea or vomiting (59.7%), and abdominal pain (56.9%) (Fig 2B and S2 Fig). In bivariable analyses, headache, haemorrhage, sore throat, diarrhoea, and myalgia were significantly associated with GTOV infection (*p* < 0.05) (S4 Table and S2 Fig).

In multivariable Poisson regression models, headache, sore throat, and diarrhoea remained independently associated with GTOV infection (RR > 1.0; *p*< 0.05), indicating a distinct clinical profile relative to comparator infections (S3 Fig).

### Case fatality ratio

From January 2017 to December 2024, a total of 72 deaths were recorded among suspected VHF cases, corresponding to an overall case fatality ratio of 15.0% (95% CI: 11.9–18.5). Among individuals with laboratory-confirmed VHF, 26 deaths occurred, corresponding to a markedly higher case fatality ratio of 36.1% (95% CI: 25.1–48.3). VHF-related deaths were predominantly observed in males (n =18; 69.2%; 95% CI: 48.2–85.7) and in individuals aged 46–90 years (n = 10; 38.5%; 95% CI: 20.2–59.4).

Among fatal VHF cases, the most frequently reported symptoms included fever (100.0%), headache (96.2%), myalgia (92.3%), arthralgia (80.8%), chills (76.9%), haemorrhage (65.4%), nausea or vomiting (65.4%), and abdominal pain (50.0%), consistent with a clinically severe presentation (S2 Fig).

GTOV infection was significantly associated with mortality. In bivariable models, GTOV infection was associated with an increased risk of death (*P* < 0.05). This association remained significant after adjustment for age and sex in multivariable Poisson regression, with a 3.66-fold higher risk of death (95% CI: 2.28–5.87; *p* < 0.001) (S3 Table).

### Clinical timeline and outcome patterns in patients with VHF

Patients presented for initial medical assessment a median of 4 days after symptom onset (IQR: 2–6). Hospital admission occured by day 5 (IQR: 3–7), and clinical specimens were collected on day 6 (IQR: 4–8). Among fatal cases, the median interval from symptom onset to death was 11 days (IQR: 8–14) (S4 Fig).

### Comparative clinical profiling of GTOV and other endemic arboviral infections

Compared with encephalitic alphaviruses (MADV and VEEV), GTOV infection was more likely to present with myalgia, haemorrhage, and sore throat. Compared with CHIKV, GTOV infection was more frequently associated with headache, retro-orbital pain, and nausea or vomiting. Compared with DENV, GTOV infection was associated with higher risks of headache, arthralgia, haemorrhage, sore throat, and respiratory symptoms. Compared with ZIKV, GTOV infection was more strongly associated with headache, haemorrhage, and abdominal pain (Fig 3).

**Fig 3.**
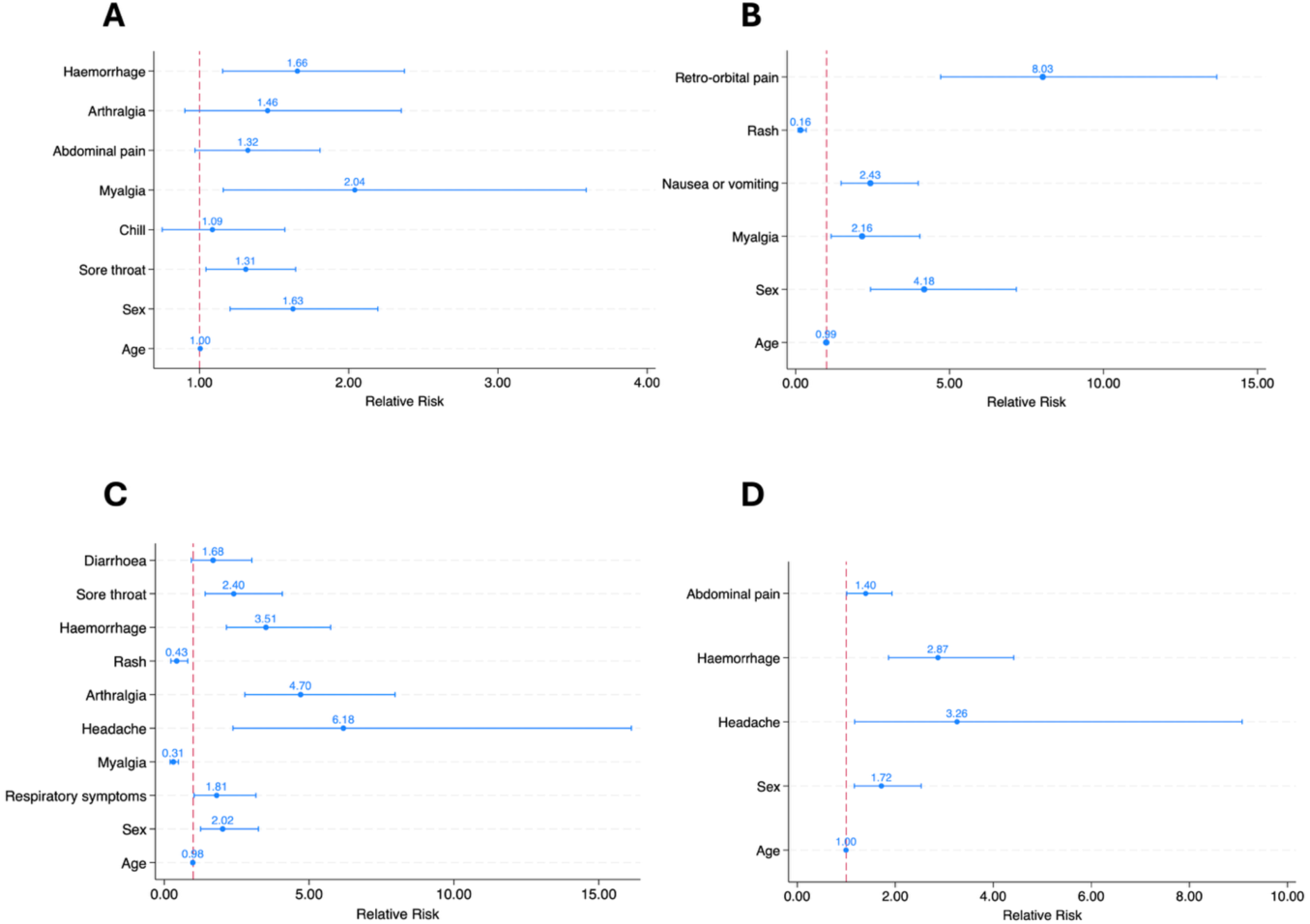
Multivariable Poisson regression analysis of symptoms associated with GTOV compared to endemic arboviruses in Venezuela. (A) GTOV compared with encephalitic alphaviruses, including Madariaga virus (MADV) and Venezuelan equine encephalitis virus (VEEV). (B) GTOV compared with chikungunya virus (CHIKV). (C) GTOV compared with dengue virus (DENV). (D) GTOV compared with Zika virus (ZIKV). Points represent relative risk (RR), and horizontal lines indicate 95% confidence intervals. The vertical red line corresponds to RR = 1.0, indicating no difference in risk between groups. All final models were adjusted for sex and age.

## Discussion

Our findings reinforce that laboratory-confirmed GTOV infections occur predominantly among males engaged in agriculture, livestock, and service-related activities, in keeping with prior evidence linking occupational exposure to rodent reservoirs [12,17]. However, the occurrence of infections among women and individuals outside traditionally rural occupations suggests that transmission may also occur in peridomestic settings, consistent with earlier investigations that isolated GTOV from rodents in close proximity to households [6]. These observations underscore the complexity of exposure pathways and suggest that risk may extend beyond strictly occupational environments.

Contrary to earlier reports identifying young adults (20–30 years) as the most affected group, we observed the highest prevalence among older adults (46–90 years), potentially reflecting age-related vulnerability [18]. Nevertheless, the multivariable analyses did not demonstrate statistically significant associations with sex, age, or occupation. These results should be interpreted with caution, as a lack of statistical significance does not necessarily indicate the absence of an effect. These findings highlight the need for larger, prospective studies to disentangle demographic and occupational risks.

Our data confirm the continued endemic circulation of GTOV in the western plains of Venezuela, with confirmed cases in Barinas, Portuguesa, and Apure—regions historically recognised as endemic or high-risk areas [19,20]. The identification of cases in Apure during the surveillance period may suggest geographical expansion or improved detection. The observed seasonal peak between December and January coincides with periods of intensified agricultural activity and potentially increased human–rodent contact, a pattern consistent with previous epidemiological observations linking VHF occurence to agricultural cycles in endemic regions [7,21].

Clinically, VHF presented as an acute febrile syndrome that, in some cases, progressed to haemorrhage and death. Laboratory-confirmed GTOV infections were more strongly associated with headache, haemorrhage, sore throat, diarrhoea, and myalgia than probable cases. Although the overall case fatality ratio (15.0%) was lower than historical estimates (23–33%), this likely reflects broader surveillance inclusion and earlier clinical recognition [6,19]. When analyses were restricted to laboratory-confirmed cases, the case fatality ratio increased to 36.1%, emphasising the severity of confirmed infections and suggesting that broader clinical case definitions may dilute estimates of lethality. The strong association between GTOV infection and mortality further reinforces the substantial pathogenic potential of this virus.

Importantly, although VHF shares substantial clinical overlap with other endemic arboviral infections, our comparative analyses identified several distinguishing features. GTOV infection was more frequently associated with haemorrhage, sore throat, and myalgia than neurotropic alphaviruses; with retro-orbital pain, nausea, and headache compared with CHIKV; and with higher frequencies of headache, arthralgia, haemorrhage, and respiratory symptoms relative to DENV. Compared with ZIKV, GTOV showed stronger associations with headache, haemorrhage, and abdominal pain. Although these features are not pathognomonic, they may aid clinical suspicion in remote endemic settings where laboratory diagnostic capacity remains limited, a persistent challenge highlighted in previous reports of misclassification and delayed diagnosis.

Several limitations merit consideration. Reliance on passive surveillance may have led to an underestimation of mild or subclinical infections, and incomplete laboratory testing reduced the sample size and representativeness. The limited number of confirmed cases constrained statistical power, and the absence of detailed data on comorbidities, exposure histories, and ecological variables restricted confounder adjustment for potential cofounders. As with all retrospective analyses of surveillance data, data completeness and reporting quality may have influenced the estimates.

Despite these limitations, this study provides the most comprehensive contemporary epidemiological and clinical characterisation of VHF in western Venezuela. Our findings underscore the need to strengthen laboratory diagnostic capacity, enhance active eco-epidemiological surveillance, and integrate zoonotic monitoring into public health response frameworks. Future investigations should prioritise the identification of subclinical infections, characterisation of peridomestic transmission pathways, and evaluation of socio-environmental drivers that shape human exposure risk in endemic regions.

## Conclusion

This study demonstrates the continued circulation of GTOV in Venezuela’s western plains, characterised by pronounced seasonal peaks. Although the majority of cases occurred among agricultural workers, infections in women and individuals outside rural occupations suggest transmission pathways extending beyond strictly occupational exposure. Clinically, we identified a distinctive symptom profile associated with GTOV infection, including higher frequencies of haemorrhage, abdominal pain, and sore throat compared with other co-circulating arboviruses. Notably, the case fatality ratio more than doubled when the analysis was restricted to laboratory-confirmed cases, underscoring the greater severity of confirmed infections and suggesting that reliance on broader suspected case definitions may underestimate the true lethality of the disease.

## Data Availability

Due to data privacy considerations and a data use agreement with the Ministry of People Power for Health (Ministerio del Poder Popular para la Salud), Barinas, Venezuela, the data cannot be made publicly available. Data may be available upon reasonable request to the Ministry of People Power for Health, subject to approval and in accordance with national and institutional regulations governing the use of surveillance data.

## Author contributions

**Conceptualisation:** María-Mercedes García, José Narciso Franco.

**Methodology:** María-Mercedes García, José Narciso Franco, Xacdiel Rodríguez, Jean-Paul Carrera.

**Data curation:** Cassia F. Estofolete, Carlos Lezcano-Coba, Josefrancisco Galué, Yelissa Juárez.

**Formal analysis:** Xacdiel Rodríguez.

**Validation:** Xacdiel Rodríguez.

**Investigation:** María-Mercedes García, Santiago José López, José José Reyes Dorante, Emilsa Jaqueline Aldana, Niyuris Elena Orduño, Aurora Lugo, Daniel Salazar, Nailyn Carvallo, Yismar Rivas, José Narciso Franco.

**Resources:** Cassia F. Estofolete, Mauricio Lacerda Nogueira, Carlos Lezcano-Coba, Josefrancisco Galué, Yelissa Juárez.

**Project administration:** María-Mercedes García, José Narciso Franco.

**Supervision:** María-Mercedes García, José Narciso Franco, Jean-Paul Carrera.

**Visualisation:** Xacdiel Rodríguez.

**Writing – original draft:** Xacdiel Rodríguez.

**Writing – review & editing:** Jean-Paul Carrera, Christl A. Donnelly, Mauricio Lacerda Nogueira.

## Data Sharing statement

Due to data privacy considerations and a data use agreement with the Ministry of People’s Power for Health (Ministerio del Poder Popular para la Salud), Barinas, Venezuela, the data cannot be made publicly available.

## Acknowledgments

The authors would like to thank the Ministry of People’s Power for Health (Ministerio del Poder Popular para la Salud), Barinas, Venezuela, for providing access to the Venezuelan haemorrhagic fever (VHF) surveillance data used in this analysis. We also extend our sincere appreciation to Mr Alberto Cumbrera for his valuable technical support in the design and preparation of the spatial maps.

## Supplementary Materials

**S1 Fig.**
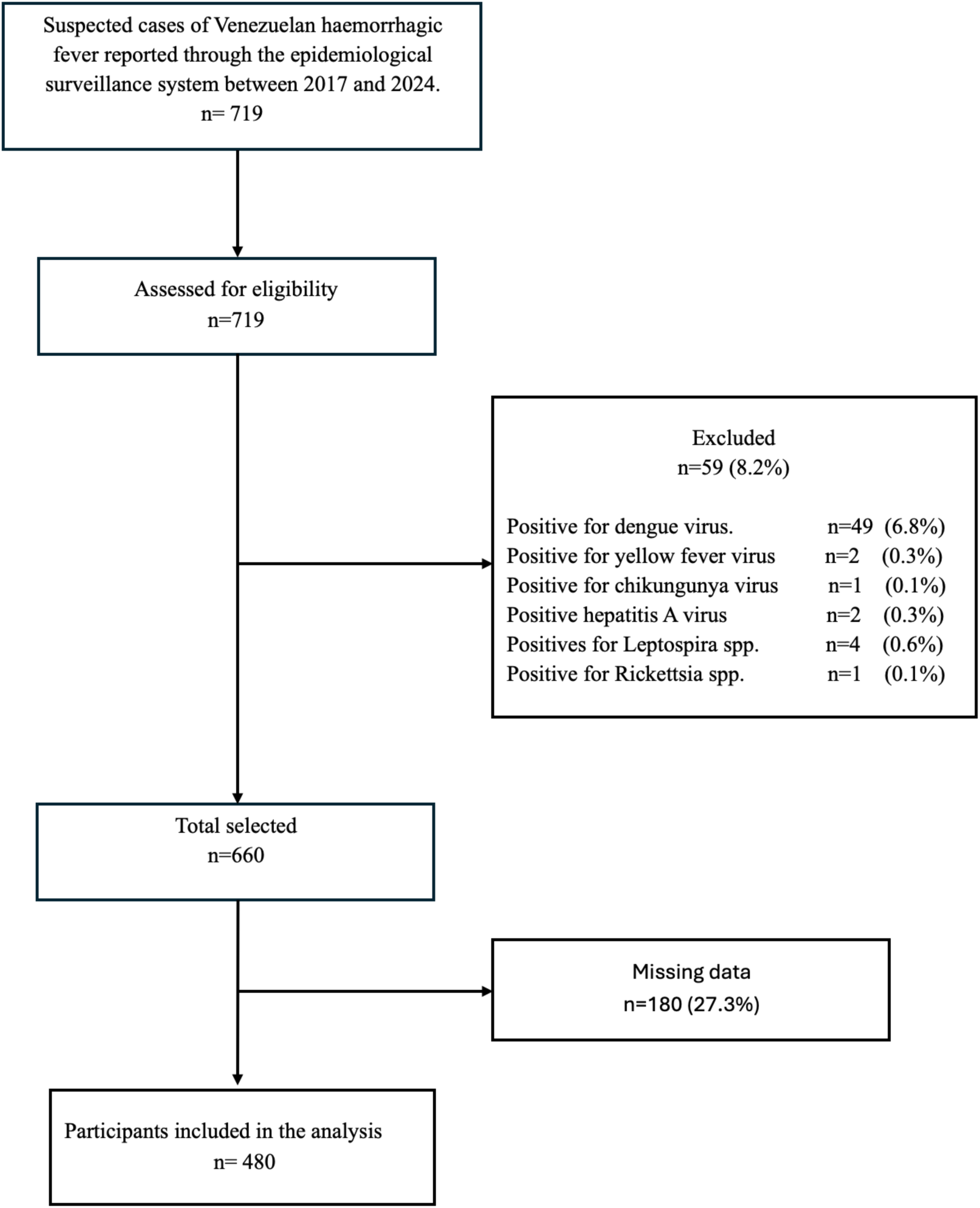
Flowchart of participant selection for the analysis of Venezuelan haemorrhagic fever cases reported through the national surveillance system, 2017–2024. A total of 719 suspected VHF cases were initially reported. Of these, 59 cases (8.2%) were excluded due to laboratory confirmation of alternative etiologies, including dengue virus, yellow fever virus, chikungunya virus, hepatitis A virus, *Leptospira* spp., and *Rickettsia* spp. After exclusions, 660 cases remained, from which 180 were removed due to missing data (27.3%). The final analytic sample included 480 participants.

**S1 Table.** Demographic and epidemiological characteristics of the study population (n= 480).

| Characteristics |  | N (%) |
| --- | --- | --- |
| Sex |  |  |
|  | Female | 155 (32.4) |
|  | Male | 324 (67.6) |
| Age (years)* |  | 29.7 ± 18.6 |
| Age (quintiles) |  |  |
|  | Q1: 1-13 years | 97 (20.2) |
|  | Q2: 14-22 years | 99 (20.6) |
|  | Q3: 23-31 years | 100 (20.8) |
|  | Q4: 32-45 years | 89 (18.5) |
|  | Q5: 46-90 years | 95 (19.8) |
| Sector of occupation |  |  |
|  | Not economically active | 105 (21.9) |
|  | Educational sector | 146 (30.4) |
|  | Agricultural and livestock sector | 156 (32.5) |
|  | Security sector | 4 (0.8) |
|  | Trade and services sector | 65 (13.5) |
|  | Health services | 4 (0.8) |
| Rodents around the house† |  |  |
|  | No | 20 (4.2) |
|  | Yes | 456 (95.8) |
| Crops near the house† |  |  |
|  | No | 16 (3.4) |
|  | Yes | 458 (96.6) |
| Use of footwear† |  |  |
|  | No | 41 (8.7) |
|  | Yes | 430 (91.3) |
| Deaths† |  |  |
|  | No | 407 (85.0) |
|  | Yes | 72 (15.0) |
| Guanarito infection confirmed |  |  |
|  | No | 408 (85.0) |
|  | Yes | 72 (15.0) |

† Some variables add up to less than 480 due to missing data.
\* Mean ± standard deviation.

**S2 Table.** Bivariate analysis of characteristics associated with Guanarito infection, (n= 480).

| Characteristics |  | Guanarito infection |  | p<sup>a</sup> |
| --- | --- | --- | --- | --- |
|  |  | No (n =408) (n %) | Yes (n =72) (n %) |  |
| Sex |  |  |  | 0.148 |
|  | Female | 137 (88.4) | 18 (11.6) |  |
|  | Male | 270 (83.3) | 54 (16.7) |  |
| Age (years)* |  | 29.3 ± 18.4 | 32.7 ± 19.8 | 0.182<sup>b</sup> |
| Age (quintiles) |  |  |  | 0.623 |
|  | Q1: 1-13 years | 84 (86.6) | 13 (13.4) |  |
|  | Q2: 14-22 years | 84 (85.8) | 15 (15.2) |  |
|  | Q3: 23-31 years | 86 (86.0) | 14 (14.0) |  |
|  | Q4: 32-45 years | 78 (87.6) | 11 (12.4) |  |
|  | Q5: 46-90 years | 76 (80.0) | 19 (20.0) |  |
| Sector of occupation |  |  |  | 0.650<sup>c</sup> |
|  | Not economically active | 94 (89.5) | 11(10.5) |  |
|  | Educational Sector | 127 (87.0) | 19 (13.1) |  |
|  | Agricultural and livestock sector | 128 (82.0) | 28 (18.0) |  |
|  | Security sector | 4 (100.0) | 0 (0.0) |  |
|  | Trade and services sector | 51 (78.5) | 14 (21.5) |  |
|  | Health services | 4 (100.0) | 0 (0.0) |  |
| Rodents around the house† |  |  |  | 0.184 |
|  | No | 15 (75.0) | 5 (25.0) |  |
|  | Yes | 391 (85.8) | 65 (14.2) |  |
| Crops near the house† |  |  |  | 0.152 |
|  | No | 12 (75.0) | 4 (25.0) |  |
|  | Yes | 393 (85.8) | 65 (14.2) |  |
| Use of footwear† |  |  |  | 0.228 |
|  | No | 32 (78.0) | 9 (22.0) |  |
|  | Yes | 371 (86.3) | 59 (13.7) |  |
| Deaths† |  |  |  | <0.001 |
|  | No | 362 (88.9) | 45 (11.1) |  |
|  | Yes | 46 (63.9) | 26 (36.1) |  |

† Some variables add up to less than 480 due to missing data.
\* Mean ± standard deviation.
<sup>a</sup> Chi square (Chi2) p-value.
<sup>b</sup> Student's t p-value
<sup>c</sup> Fisher's exact p-value

**S3 Table.** Characteristics associated with Guanarito infection, regression analysis (n= 480).

| <u>Variables</u> | <u>Bivariate Analysis</u> |  |  | <u>Multivariable Analysis</u> |  |  |
| --- | --- | --- | --- | --- | --- | --- |
|  | RR | IC 95% | p* | RR | IC 95% | p** |
| Sex |  |  |  |  |  |  |
| Female | Ref. |  |  | Ref. |  |  |
| Male | 1.44 | 0.87 – 2.36 | 0.155 | 0.82 | 0.41 – 1.67 | 0.593 |
| Age (quintiles) |  |  |  |  |  |  |
| Q1: 1-13 years | Ref. |  |  | Ref. |  |  |
| Q2: 14-22 years | 1.13 | 0.56 – 2.25 | 0.727 | 0.90 | 0.43 – 1.89 | 0.793 |
| Q3: 23-31 years | 1.04 | 0.52 – 2.11 | 0.903 | 0.75 | 0.32 – 1.74 | 0.501 |
| Q4: 32-45 years | 0.92 | 0.44 – 1.95 | 0.832 | 0.69 | 0.29 – 1.64 | 0.405 |
| Q5: 46-90 years | 1.49 | 0.78 – 2.85 | 0.225 | 0.64 | 0.27 – 1.51 | 0.308 |
| Sector of occupation |  |  |  |  |  |  |
| Not economically active | Ref. |  |  | Ref. |  |  |
| Educational Sector | 1.24 | 0.62 – 2.50 | 0.543 | 1.24 | 0.59 – 2.60 | 0.569 |
| Agricultural and livestock sector | 1.71 | 0.89 – 3.29 | 0.106 | 2.03 | 0.87 – 4.73 | 0.101 |
| Trade and services sector | 2.06 | 0.99 – 4.25 | 0.052 | 2.21 | 0.85 – 5.76 | 0.103 |
| Rodents around the house† |  |  |  |  |  |  |
| No | Ref. |  |  | Ref. |  |  |
| Yes | 0.58 | 0.26 – 1.28 | 0.178 | 0.97 | 0.19 – 5.00 | 0.974 |
| Crops near the house† |  |  |  |  |  |  |
| No | Ref. |  |  | Ref. |  |  |
| Yes | 0.57 | 0.24 – 1.37 | 0.207 | 1.57 | 0.38 – 6.55 | 0.523 |
| Use of footwear† |  |  |  |  |  |  |
| No | Ref. |  |  | Ref. |  |  |
| Yes | 0.64 | 0.34 – 1.19 | 0.157 | 0.54 | 0.18 – 1.62 | 0.272 |
| Death† |  |  |  |  |  |  |
| No | Ref. |  |  | Ref. |  |  |
| Yes | 3.20 | 2.12 – 4.84 | <0.001 | 3.66 | 2.28 – 5.87 | <0.001 |

† Some variables add up to less than 480 due to missing data.
\* *p*-value obtained from the generalised linear model (GLM, Poisson family, log-link function, with robust variance adjustment) for the bivariate association between Guanarito virus infection and covariates.
\*\* *p*-value obtained from the generalised linear model (GLM, Poisson family, log-link function, with robust variance adjustment) for the association between Guanarito virus infection and covariates, adjusted for sex and age.

**S4 Table.**
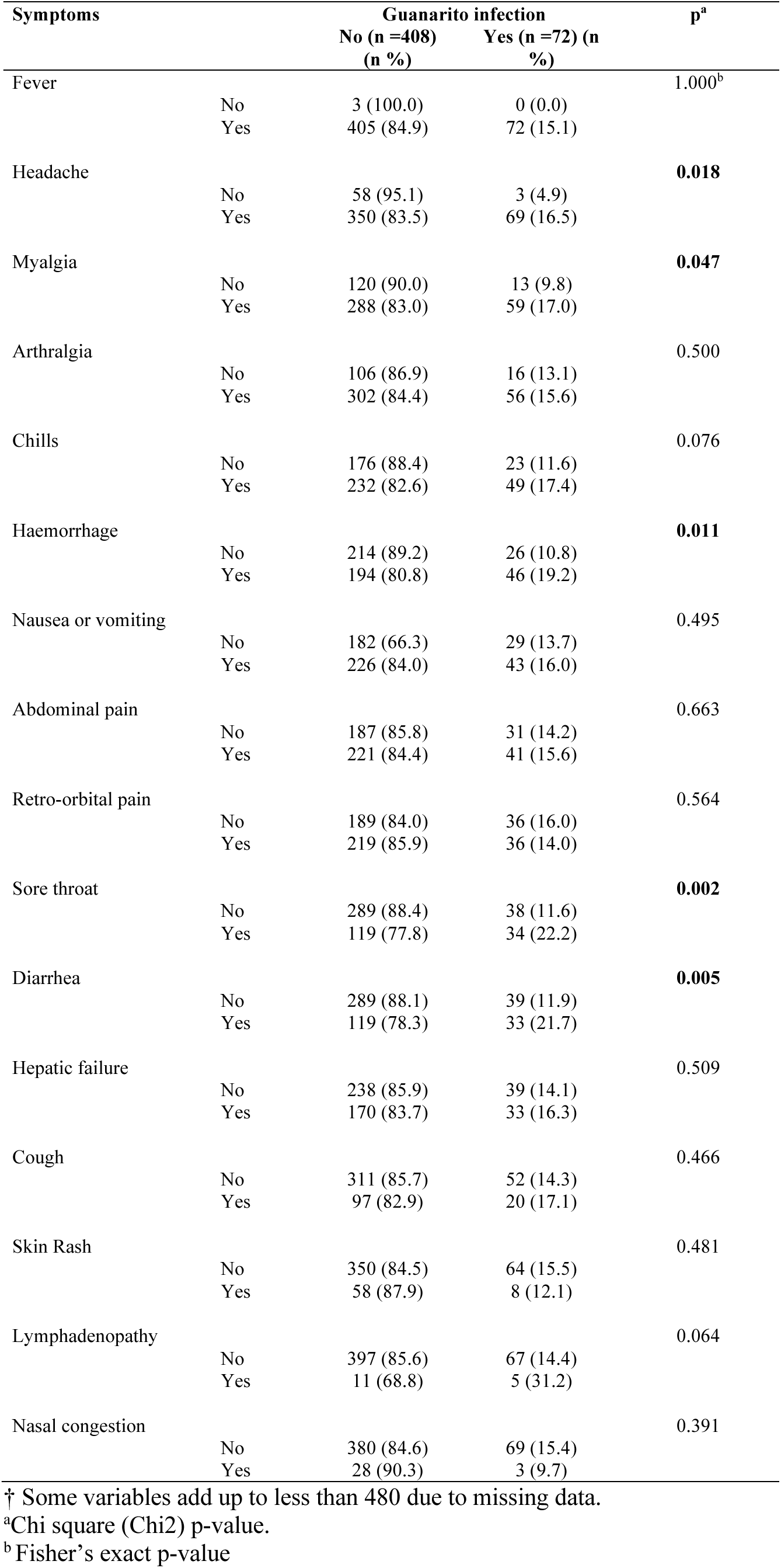
Frequency and bivariate analysis of signs and symptoms.

| Symptoms |  | Guanarito infection |  | p<sup>a</sup> |
| --- | --- | --- | --- | --- |
|  |  | No (n =408)<br>(n %) | Yes (n =72) (n<br>%) |  |
| Fever | No | 3 (100.0) | 0 (0.0) | 1.000<sup>b</sup> |
|  | Yes | 405 (84.9) | 72 (15.1) |  |
| Headache | No | 58 (95.1) | 3 (4.9) | **0.018** |
|  | Yes | 350 (83.5) | 69 (16.5) |  |
| Myalgia | No | 120 (90.0) | 13 (9.8) | **0.047** |
|  | Yes | 288 (83.0) | 59 (17.0) |  |
| Arthralgia | No | 106 (86.9) | 16 (13.1) | 0.500 |
|  | Yes | 302 (84.4) | 56 (15.6) |  |
| Chills | No | 176 (88.4) | 23 (11.6) | 0.076 |
|  | Yes | 232 (82.6) | 49 (17.4) |  |
| Haemorrhage | No | 214 (89.2) | 26 (10.8) | **0.011** |
|  | Yes | 194 (80.8) | 46 (19.2) |  |
| Nausea or vomiting | No | 182 (66.3) | 29 (13.7) | 0.495 |
|  | Yes | 226 (84.0) | 43 (16.0) |  |
| Abdominal pain | No | 187 (85.8) | 31 (14.2) | 0.663 |
|  | Yes | 221 (84.4) | 41 (15.6) |  |
| Retro-orbital pain | No | 189 (84.0) | 36 (16.0) | 0.564 |
|  | Yes | 219 (85.9) | 36 (14.0) |  |
| Sore throat | No | 289 (88.4) | 38 (11.6) | **0.002** |
|  | Yes | 119 (77.8) | 34 (22.2) |  |
| Diarrhea | No | 289 (88.1) | 39 (11.9) | **0.005** |
|  | Yes | 119 (78.3) | 33 (21.7) |  |
| Hepatic failure | No | 238 (85.9) | 39 (14.1) | 0.509 |
|  | Yes | 170 (83.7) | 33 (16.3) |  |
| Cough | No | 311 (85.7) | 52 (14.3) | 0.466 |
|  | Yes | 97 (82.9) | 20 (17.1) |  |
| Skin Rash | No | 350 (84.5) | 64 (15.5) | 0.481 |
|  | Yes | 58 (87.9) | 8 (12.1) |  |
| Lymphadenopathy | No | 397 (85.6) | 67 (14.4) | 0.064 |
|  | Yes | 11 (68.8) | 5 (31.2) |  |
| Nasal congestion | No | 380 (84.6) | 69 (15.4) | 0.391 |
|  | Yes | 28 (90.3) | 3 (9.7) |  |

† Some variables add up to less than 480 due to missing data.
<sup>a</sup>Chi square (Chi2) p-value.
<sup>b</sup>Fisher's exact p-value

**S2 Fig.**
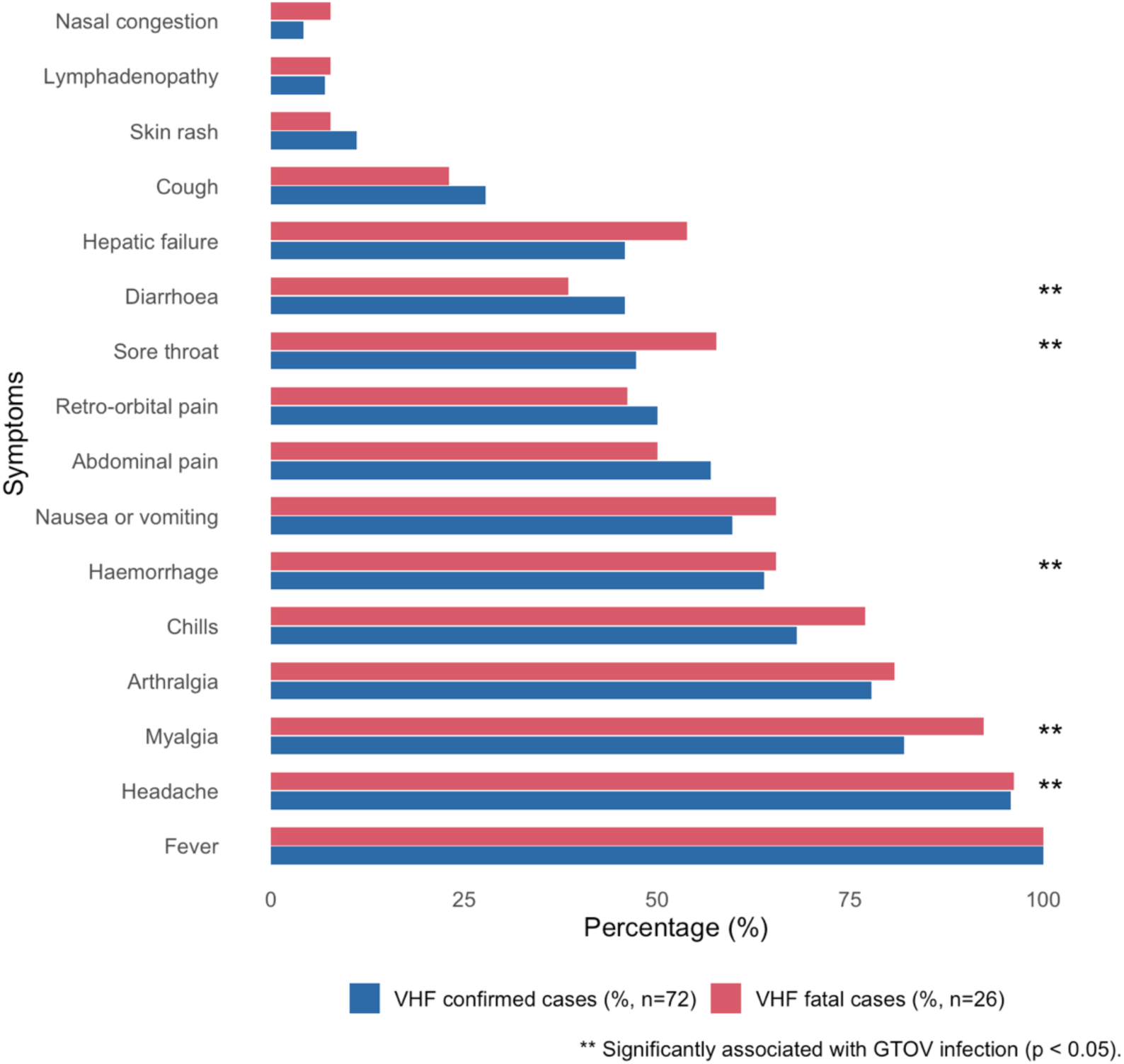
Clinical symptoms among confirmed and fatal cases of Venezuelan haemorrhagic fever. Distribution of clinical symptoms among patients with confirmed VHF. Bars represent the percentage of patients presenting each symptom among confirmed cases (blue; n = 72) and fatal cases (red; n = 26). Percentages were calculated within each group. Asterisks indicate symptoms significantly associated with GTOV infection in bivariable analysis (p < 0.05).

**S3 Fig.**
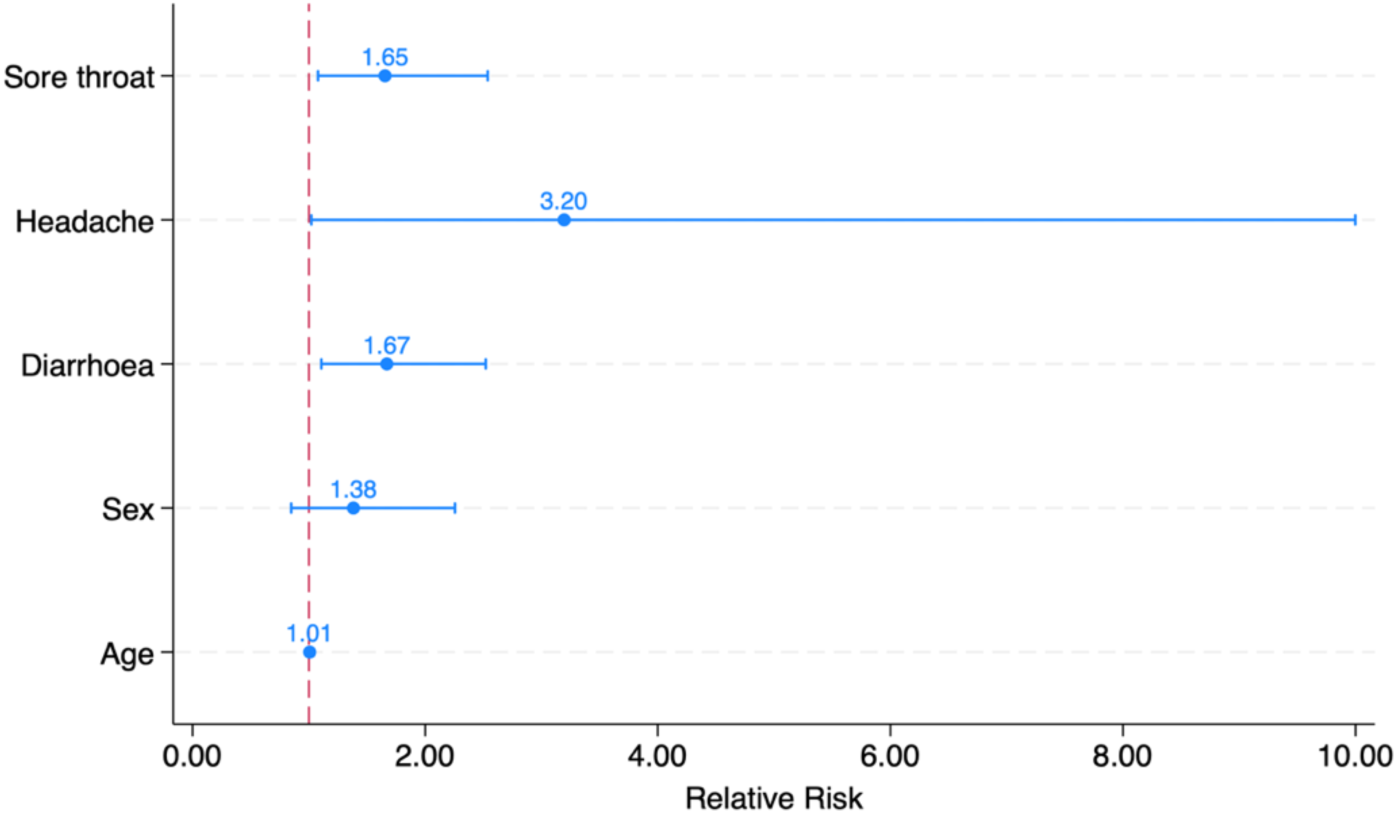
Multivariable Poisson regression analysis of factors associated with GTOV infection. Points represent adjusted relative risks (RRs), and horizontal lines indicate 95% confidence intervals. The vertical red line corresponds to an RR of 1.0, indicating no association.

**S4 Fig.**
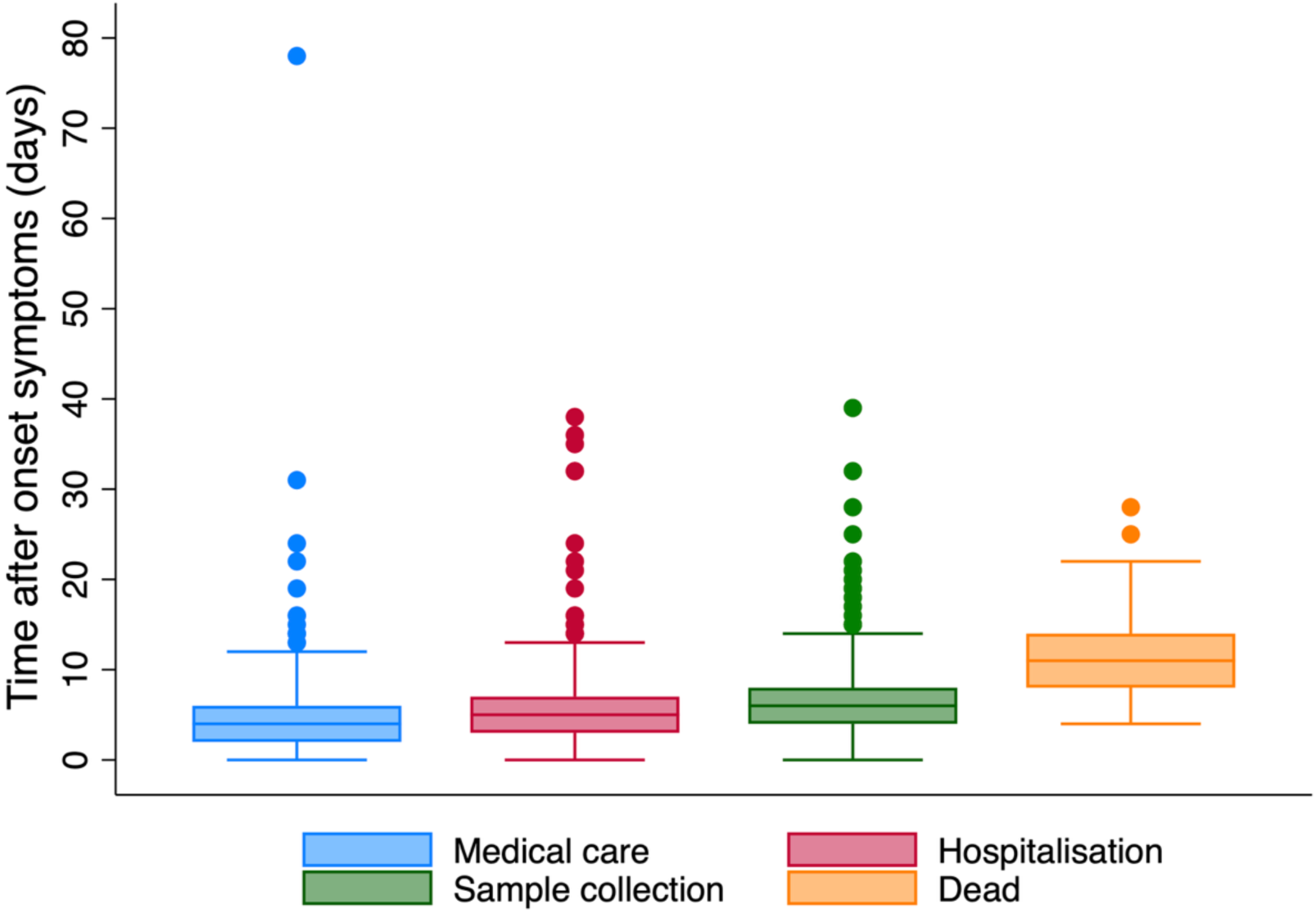
Timing of clinical events following symptom onset in confirmed cases of VHF. Boxplots showing the distribution of time (in days) between symptom onset and key clinical events among confirmed cases of VHF. Events include first medical consultation (blue), hospitalisation (red), sample collection for laboratory confirmation (green), and time of death (orange). The median, interquartile range, and outliers are represented for each category.

**S5 Table.** Venezuelan Haemorrhagic Fever case definition.

| FHV Cases | Definition |
| --- | --- |
| Suspect Case | A patient with an undetermined febrile illness residing in an endemic area for VHF (Portuguesa, Barinas, and Guárico), in a high-risk area (Apure and Cojedes), or who has visited these areas within the previous 21 days. |
| Probable Case | A patient from a rural area endemic for VHF (Portuguesa, Barinas, and Guárico), or from a high-risk area (Apure and Cojedes), or who has visited these areas within the previous 21 days, presenting with an insidious onset of fever and one or more of the following symptoms: general malaise, headache, arthralgia, myalgia, abdominal pain, dysphagia, odynophagia, vomiting, or diarrhoea, with or without haemorrhagic manifestations such as gingivorrhagia, epistaxis, petechiae, ecchymosis, etc., and laboratory findings of leukopenia and thrombocytopenia, in the absence of a presumptive bacterial cause. |
| Confirmed Case | Cases meeting the criteria for a probable case of VHF, in whom virological confirmation was established through the isolation and identification of Guanarito virus and/or partial amplification of viral RNA by RT-PCR, and/or serological diagnosis demonstrating a fourfold or greater rise in specific IgG antibody titres against Guanarito virus using indirect immunofluorescence assay (IFA) or ELISA techniques. |

**S6 Table.**
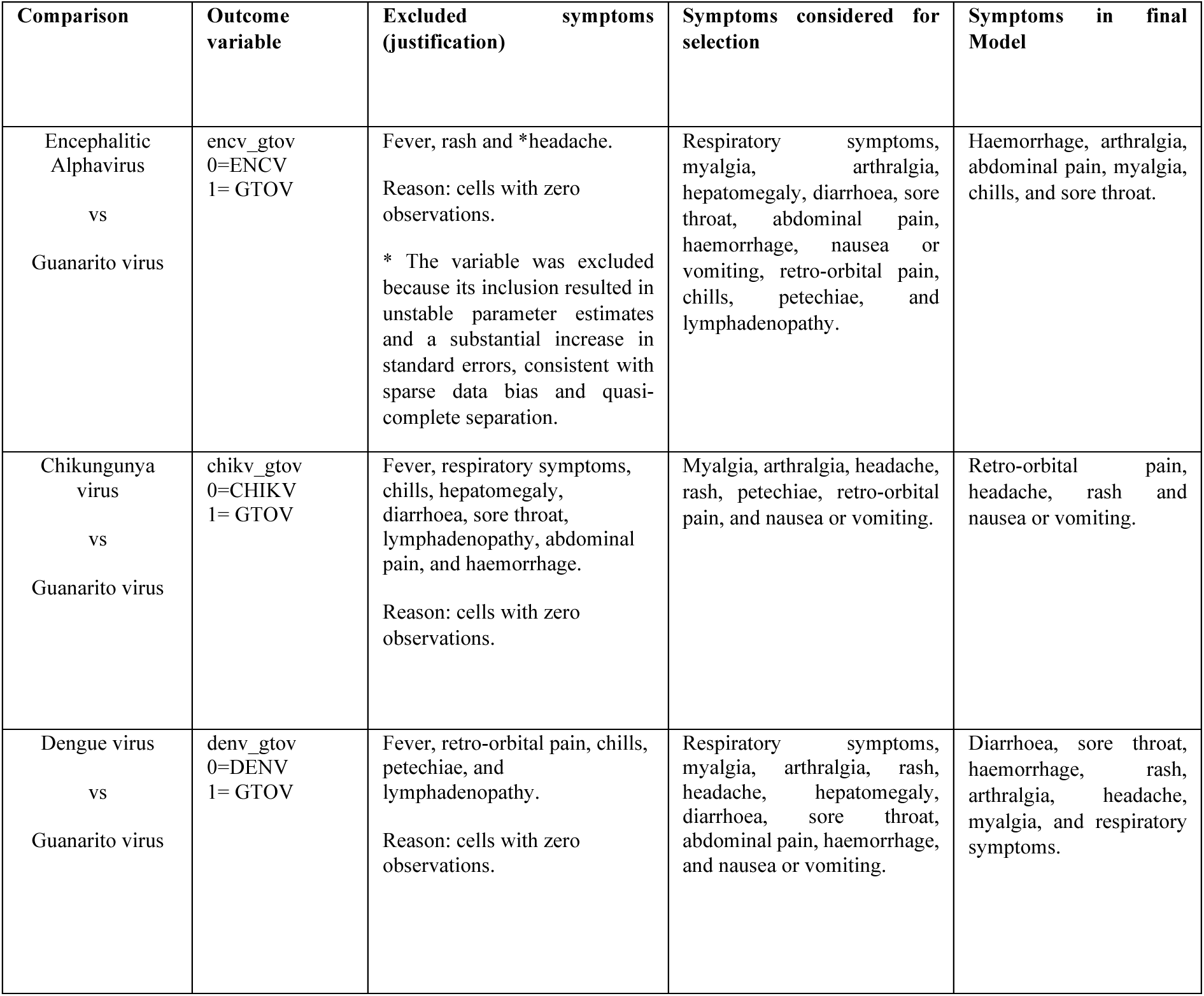
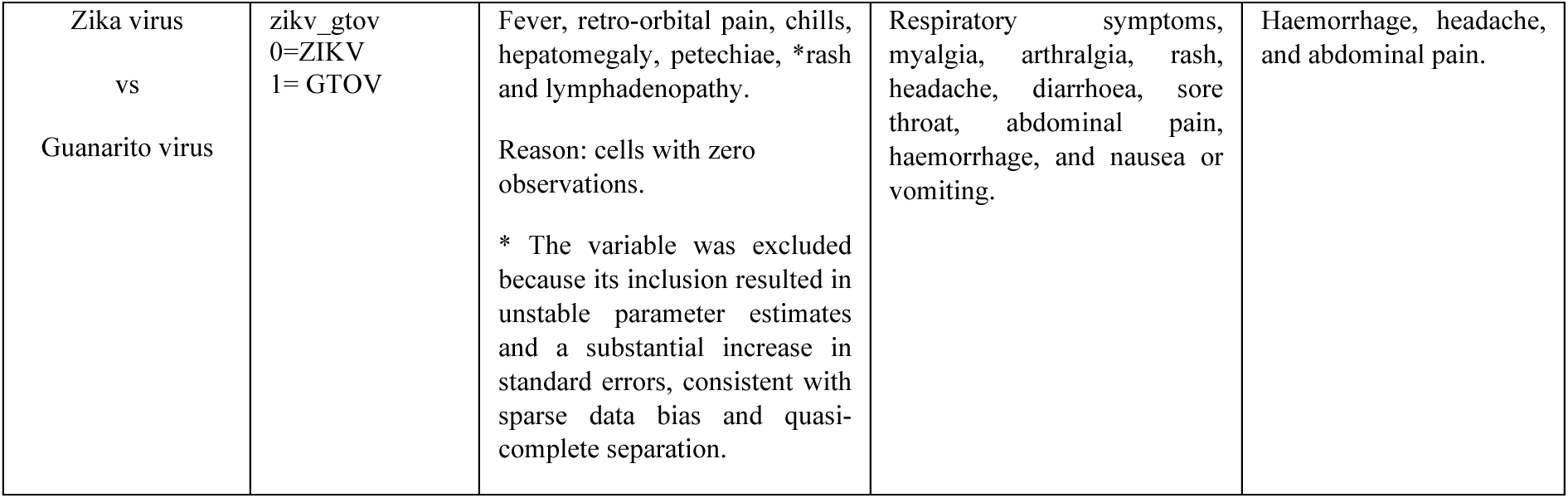
Criteria for Symptom Variable Selection and Final Inclusion in Symptom-Based Models Comparing GTOV and Other Endemic Arboviruses.

